# Reduction of Major Adverse Cardiovascular Events (MACE) after Bariatric Surgery in Obese Patients with Cardiovascular Diseases: A Systematic Review and Meta-Analysis

**DOI:** 10.1101/2021.09.16.21263439

**Authors:** Andryanto Sutanto, Henry Sutanto

## Abstract

Cardiovascular diseases (CVDs) are the leading cause of death worldwide and obesity is a major risk factor which increases morbidity and mortality of CVDs. Lifestyle modifications (e.g., diet control, physical exercise and behavioral changes) have been the first-line managements of obesity for decades. Nonetheless, when such interventions fail, pharmacotherapies and bariatric surgery are considered. Interestingly, a sudden weight loss (e.g., due to bariatric surgery) could also increase mortality (i.e., “obesity paradox”). Thus, it remains unclear whether the bariatric-surgery-associated weight reduction in patients with obesity and CVDs is beneficial for the reduction of Major Adverse Cardiovascular Events (MACE). Here, we performed a systematic literature search and meta-analysis of published studies comparing the MACE in patients with obesity and CVDs underwent bariatric surgery with control patients (no surgery). Studies’ data, including odds ratio (OR), were pooled and analyzed in a meta-analysis using a random effect model. Ten studies with a total of 1,772,305 patients, consisted of 74,042 patients underwent any form of bariatric surgery and 1,698,263 patients with no-surgery, were included in the meta-analysis. A random effect model was employed for analysis and showed that bariatric surgery group had significantly lower odds of MACE as compared to no surgery (OR = 0.49; 95% CI 0.40-0.60; *p*<0.00001; *I*^*2*^=93%), suggesting the benefit of bariatric surgery in reducing the occurrence of MACE in obese patients with CVDs.

## 1. INTRODUCTION

The global prevalence of cardiovascular diseases (CVDs) had doubled from 271 million cases in 1990 to 523 million cases in 2019, with mortality reaching 18.6 million cases worldwide (1), and these numbers are projected to increase in the next few years. Among all CVDs, ischemic heart disease and cerebrovascular diseases (e.g., stroke) are the major contributors to the high CVD burden. There have been a 120-137% increase of ischemic heart disease-related mortality and a 107-124% increase of stroke-related mortality in the past two decades (2). Such a marked increase of CVD incidence is believed to be precipitated by several factors, including population aging, urbanization and technological advancement, which lead to sedentary lifestyle and obesity, two of four canonical risk factors of CVDs, together with tobacco smoking and unhealthy diet (1, 2).

Obesity (a body mass index [BMI] more than 30 kg/m^2^ or 28 kg/m^2^ in Asian population) is known to modulate the risk for developing ischemic heart disease, cardiac arrhythmias and heart failure (HF) through several mechanisms. The excess of adipose tissue leads to insulin resistance, inflammation, the activation of renin-angiotensin-aldosterone system (RAAS), and the progressive structural and electromechanical remodeling of the heart (3, 4). Therefore, reducing the accumulation of adipose tissue is essential for obese individuals to lower the CVD risk and burden. Lifestyle modifications (e.g., diet control, physical exercise and behavioral changes) are the first-line managements of obesity (5). However, when these interventions fail to significantly lower the BMI, pharmacotherapies and bariatric surgery are considered (6).

Bariatric surgery is by definition a surgical procedure to promote weight loss. The approach is performed by restricting gastric size to reduce the amount of food ingested and/or to facilitate malabsorption of nutrients. There are several common procedures in bariatric surgery, such as gastric banding, vertical banded gastroplasty, sleeve gastrectomy, Roux-en-Y gastric bypass and biliopancreatic diversion with/without duodenal switch (6, 7). Gastric banding (**Figure 1A**) is generally performed by placing a band around stomach to restrict gastric size, whereas in the vertical banded gastroplasty (**Figure 1B**), the stomach was partitioned and a prosthetic is placed around the partitioned stomach. Meanwhile, a sleeve gastrectomy is done by resecting part of the gastric body, creating a gastric sleeve which restricts gastric size and promotes malabsorption (**Figure 1C**). In Roux-en-Y gastric bypass, stomach is partitioned into proximal and distal parts. The proximal part acts as an alimentary tract and is anastomosed with jejunum (i.e., gastrojejunostomy), while the distal part acts as a biliopancreatic limb, which is anastomosed with jejunum in either end-to-side or side-to-side fashion (**Figure 1D**). Finally, a biliopancreatic diversion approach works quite similarly to Roux-en-Y gastric bypass by dividing stomach into alimentary and biliopancreatic limbs, although in a biliopancreatic diversion, gastric resection was also performed. Both Roux-en-Y gastric bypass and biliopancreatic diversion exert their functions by creating malabsorption (7).

**Figure 1.**
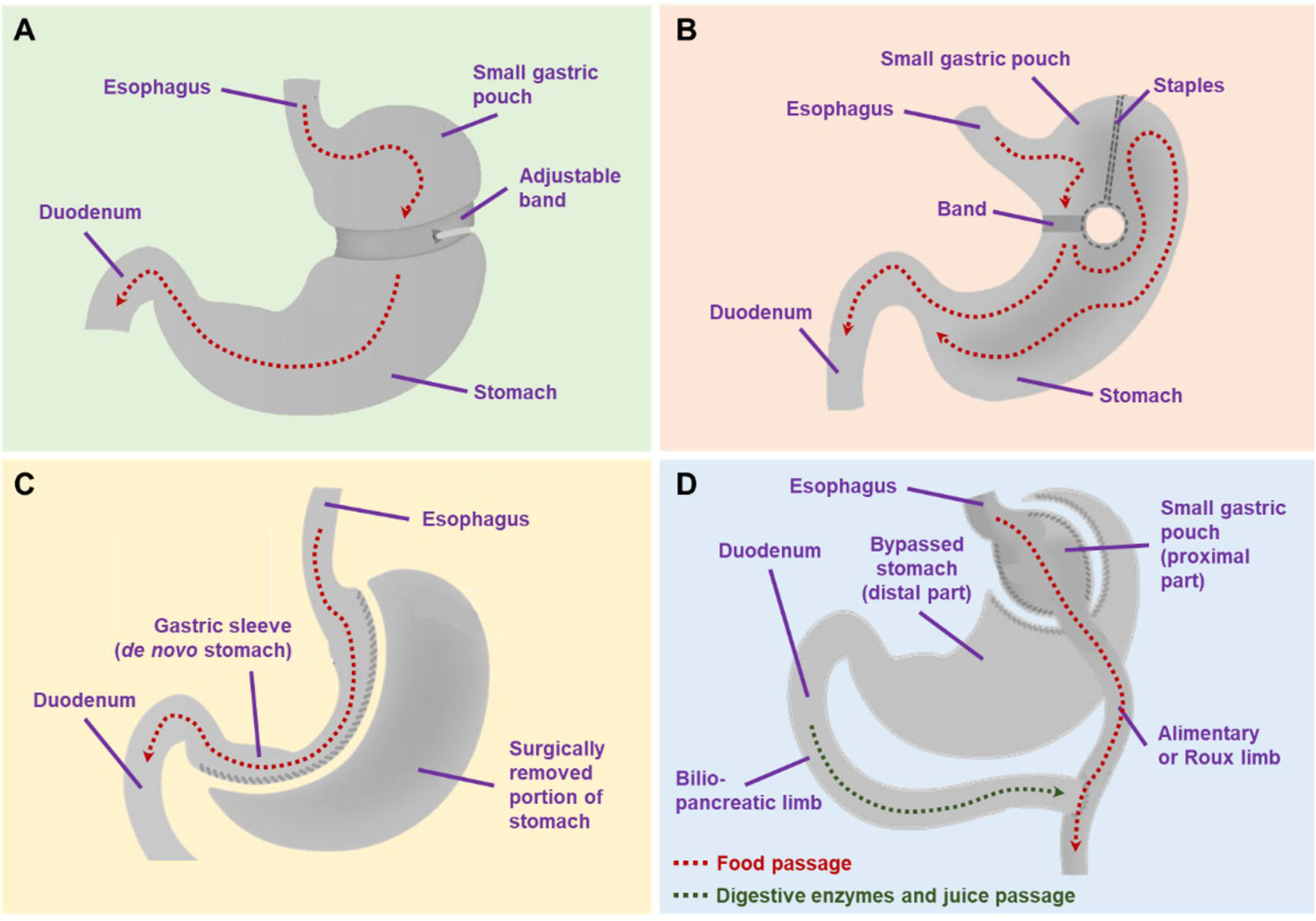
Commonly performed bariatric surgery procedures. A) gastric banding; B) vertical banded gastroplasty; C) sleeve gastrectomy; D) Roux-en-Y gastric bypass.

Several studies have reported the benefits of bariatric surgery in patients with obesity, including the improvements of body fat distribution and CV risk factors such as dyslipidemia, (pre)hypertension, insulin resistance, (pre)diabetes, non-alcoholic fatty liver disease, inflammation, vascular reactivity and obstructive sleep apnea (6). Typically, the reduction of those CV risk factors is expected to be followed by the reduction of Major Adverse Cardiovascular Events (MACE; i.e., composite of CV death, myocardial infarction [MI], stroke, coronary revascularization or hospitalization for HF (8, 9)). However, recent studies showed conflicting results, suggesting that obesity might provide better CV outcomes in specific populations (i.e., “obesity paradox”) (10-12). “Obesity paradox” is an epidemiological phenomenon in which overweight subjects or individuals with class I obesity might have better CV outcomes and/or survival, and potentially, less MACE occurrence compared to normal or underweighted subjects (3).

With the “obesity paradox” in place, it is still unknown whether a significant weight reduction would result in better CV outcomes. Moreover, at present, the benefit of bariatric surgery in reducing MACE in patients with CVDs and obesity remains poorly understood. Therefore, in this systematic review/meta-analysis, we sought to analyze the benefit of bariatric surgery in reducing MACE as compared to no surgery in patients with CVDs and obesity.

## 2. Materials and Methods

### 2.1. Search Strategy

This review was conducted in accordance with the Preferred Reporting Items for Systematic Reviews and Meta-Analysis (PRISMA) guideline (13). The PRISMA checklist is available in the ***Supplementary Materials***. Literature search was carried out electronically and relevant studies were retrieved from PubMed/MEDLINE, ScienceDirect, Cochrane Library, Wiley Online Library, and Springer databases. The search was conducted using the keywords constructed on Medical Subject Headings (MeSH) and other additional terms: “Bariatric Surgery[mesh] OR Metabolic Surgery[mesh]” AND “Cardiovascular disease*[mesh] OR Obesity” AND “Major Adverse Cardiac Event* OR Major Adverse Cardiovascular Event* OR MACE” OR “Bariatric Surgery and Long-term Cardiovascular Events”. All publications from the inception to July 2021 were evaluated.

### 2.2. Eligibility

We included studies that focused primarily on the comparison of MACE in obese patients with CVDs underwent bariatric surgery and no surgery. Studies were opted with following inclusion criteria: (1) the primary endpoint of the studies was the occurrence of MACE (defined as all-cause mortality or the first occurrence of MI, coronary artery bypass grafting or percutaneous coronary intervention, stroke, or hospitalization for HF), (2) studies comparing surgery and no-surgery groups, (3) the study population was adults with CVDs (e.g., ischemic heart disease, hypertension, HF) and obesity with an exclusion of persons with age less than 18 years old or more than 80 years old, pregnancy or malignancy, (4) the full-text of the articles is accessible, (5) randomized controlled trial (RCT) or cohort studies, and (6) the studies were published in English. Studies in the form of review articles and case reports/case series were excluded.

### 2.3. Data Collection and Extraction

Literatures were screened and reviewed by two independent reviewers (A.S. and H.S.). Screening was done by assessing the relevance of the title and abstract of the studies. Any duplication of the studies was removed using Mendeley Reference Manager. From the reference literatures, the following data were taken: the type of study design, study locations (state and/or country), number of patients studied, number of patients underwent bariatric surgery and no surgery, comorbidities (e.g., ischemic heart disease, HF, atrial fibrillation (AF), hypertension, dyslipidemia and diabetes mellitus), age of patients, BMI of patients, rate of MACE occurrence and follow-up period. The risk of bias was assessed using Cochrane Tool (14) and Newcastle-Ottawa Scale (NOS) (15) for RCTs and observational studies, respectively.

### 2.4. Data Synthesis

All outcome variables were summarized and pooled in a meta-analysis using the Review Manager (RevMan) 5.4.1 software (Cochrane Collaboration). Dichotomous data were presented by odds ratio (OR) and analyzed using Mantel-Haenszel method. Continuous data were presented in mean difference and analyzed using Inverse Variance method. Heterogeneity analysis was done with *I*^*2*^ test, and data was considered heterogenous if *I*^*2*^ >75% and in this setting, a random-effect model was used. If *I*^*2*^ <25%, data was considered homogenous and a fixed-effect model was used. Publication bias was assessed visually using the Begg’s funnel plot. In the presence of publication bias, trim and fill method was used for correction. Statistical significance was considered if the two-tailed *p*-value <0.05.

## 3. Results

### 3.1. Study Characteristics

A total of 726 studies were identified in the literature search, as depicted in the PRISMA flow diagram (**Figure 2**). Five duplicates were removed and 595 studies were excluded because of the irrelevance to the aim of this study. One hundred and twenty-six studies were thoroughly reviewed for eligibility. After a thorough review, 115 studies were excluded and the final 11 studies were included in the review. Of those, 10 studies were observational cohort studies and one study was a non-RCT. Subsequently, 10 studies were included in the meta-analysis (one study was excluded from meta-analysis due to the lack of adequate quantitative data). Risk of bias assessment was summarized in **Figure 3**.

**Figure 2.**
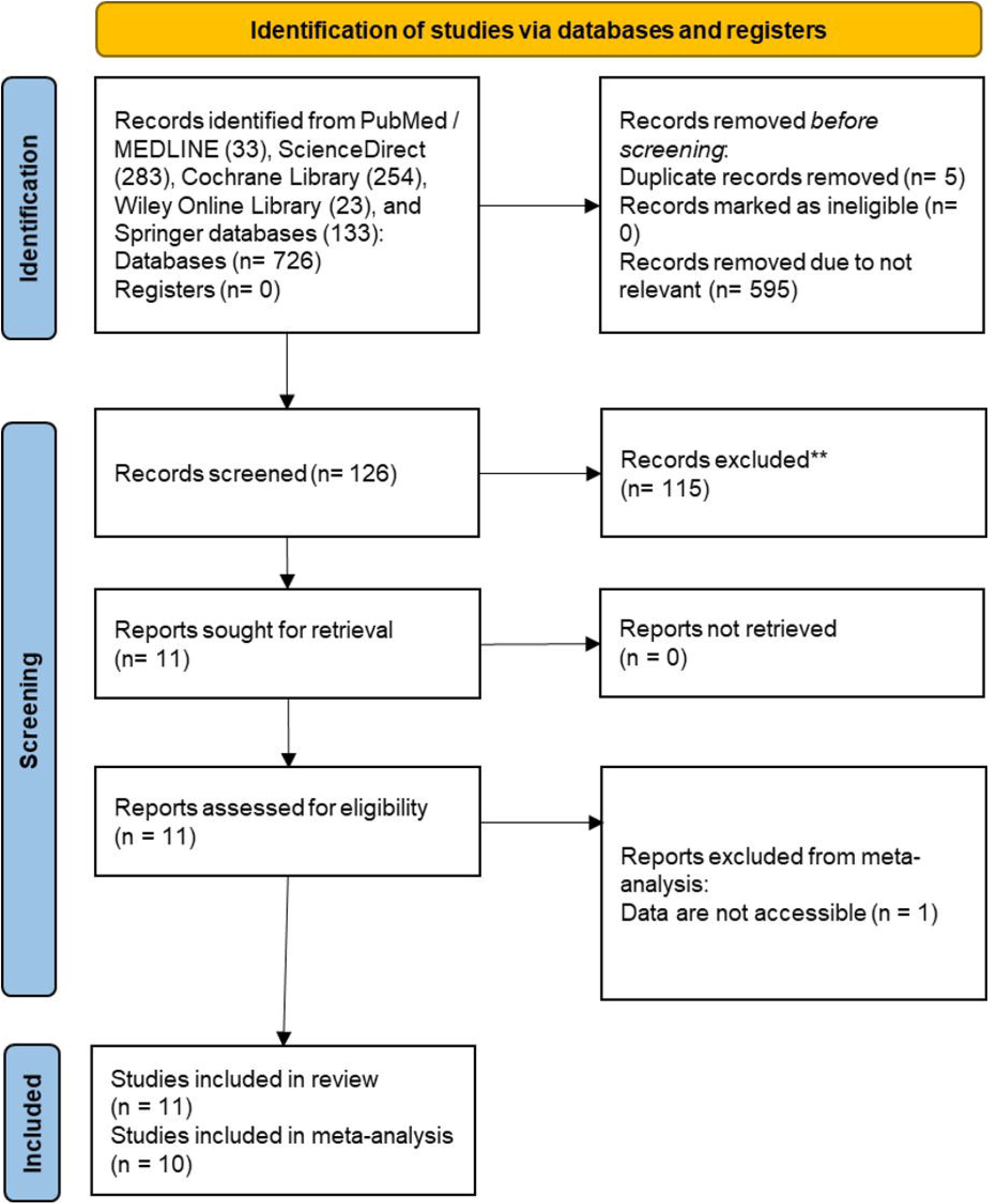
PRISMA flow diagram of literature search.

**Figure 3.**
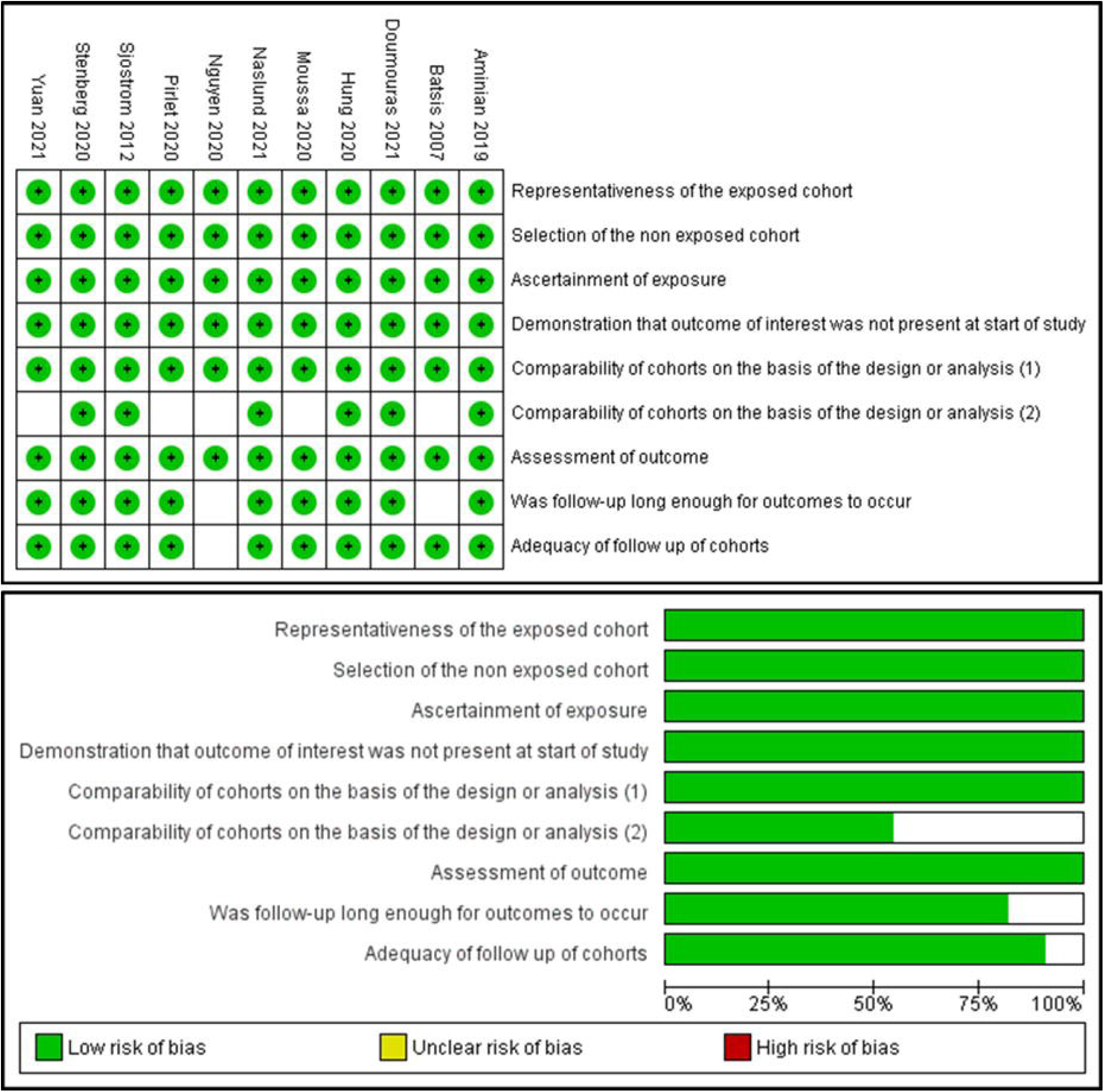
Risk of bias assessment with Newcastle-Ottawa Scale (NOS).

From 11 studies included in the review, there were 1,772,305 patients consisted of 74,042 patients underwent any form of bariatric surgery and 1,698,263 patients with no surgery. Reported bariatric procedures included Roux-en-Y gastric bypass, gastric banding, sleeve gastrectomy, biliopancreatic diversion, vertical banded gastroplasty and duodenal switch. The follow-up period of the studies ranged from 3 to 9 years. The detailed study characteristics were summarized in **Table 1**. Pooled means of age of the study population were 52.55 years in bariatric surgery group and 54.09 years in no-surgery group. Pooled means of BMI were 42.62 kg/m^2^ in bariatric surgery group and 44.59 kg/m^2^ in no-surgery group. Study population characteristics and comorbidities were listed and summarized in **Table 2**.

**Table 1.** Characteristics of the included studies.

| Study | Design | Study Location | Sample size | Surgical Procedure (%) | Follow-up Period (mean±SD / median(IQR), y) | Population | REF |
| --- | --- | --- | --- | --- | --- | --- | --- |
| Sjostrom *et al.*, 2012 | Non-RCT | Sweden | 2,010 surgery | Gastric Bypass: 265 (13.2)<br>Gastric Banding: 376 (18.7)<br>Vertical Banded Gastroplasty: 1,360 (68.1) | 14.7 | Patients aged 37 to 60 years and with BMI of at least 34 for men and at least 38 for women. | (16) |
|  |  |  | 2,037 control | - | 14.7 |  |  |
| Aminian *et al.*, 2019 | Matched Cohort Study | Florida, Ohio, USA | 2,287 surgery | Roux-en-Y Gastric Bypass: 1,443 (63)<br>Sleeve Gastrectomy: 730 (32)<br>Gastric Banding: 109 (5)<br>Duodenal Switch: 5 (0.2) | 3.3 (1.2-6.3) | Patients with age 18-80 years, BMI of 30 or greater, and diabetes. | (17) |
|  |  |  | 11,435 control |  | 4.0 (2.1-6.1) |  |  |
| Stenberg *et al.*, 2020 | Matched Cohort Study | Sweden | 11,863 surgery | Gastric Bypass: 10,692 (90.1)<br>Sleeve Gastrectomy: 1,171 (9.9) | 5.09 ± 2.53 | Patients with morbid obesity and hypertension | (18) |
|  |  |  | 26,199 control | - | 5.06 ± 2.55 |  |  |
| Pirlet *et al.*, 2020 | Matched Cohort Study | Quebec, Canada | 116 surgery | Gastric Bypass: 3 (2.6)<br>Biliopancreatic diversion with duodenal switch: 44 (38)<br>Sleeve gastrectomy: 67 (58)<br>Duodenal Switch only: 2 (1.7) | 8.9 (6.3–14.2) | Patients with history of coronary artery disease (CAD) and obesity | (19) |
|  |  |  | 116 control | - | 8.9 (6.3–14.2) |  |  |
| Moussa *et al.*, 2020 | Matched Cohort Study | UK | 3,701 surgery | N/A | 11.2 | Patients with BMI ≥ 35 kg/m<sup>2</sup> with exclusion of previous MACE | (20) |
|  |  |  | 3,701 control | - | 11.2 |  |  |
| Hung *et al.*, 2020 | Matched Cohort Study | Taiwan | 1,436 surgery | N/A | 7.5 | Patients with BMI >35 kg/m<sup>2</sup> with co-morbidities or >40 kg/m<sup>2</sup> | (22) |
|  |  |  | 1,436 control | - | 7.5 |  |  |
| Doumouras<br>*et al.*, 2021 | Matched<br>Cohort Study | Ontario,<br>Canada | 1,319<br>surgery | Gastric Bypass: 1,049<br>(79.5)<br>Sleeve Gastrectomy:<br>270 (20.5) | 4.65 (3.09–<br>6.28) | Patients with<br>BMI >35 kg/m<sup>2</sup><br>with a comorbidity<br>or BMI ≥40 kg/m<sup>2</sup> | (23) |
|  |  |  | 1,319<br>control | - | 4.38 (2.83–<br>6.09) |  |  |
| Naslund *et al.*, 2021 | Matched<br>Cohort Study | Sweden | 509<br>surgery | Roux-en-Y Gastric<br>Bypass: 465 (91)<br>Sleeve Gastrectomy:<br>44 (9) | 4.6 (2.7–7.1) | Patients with<br>severe obesity<br>and history of MI | (21) |
|  |  |  | 509<br>control | - | 4.6 (2.7–7.1) |  |  |
| Batsis *et al.*, 2007 | Retrospective<br>Cohort | Olmsted,<br>Minnesota,<br>USA | 197<br>surgery | Roux-en-Y Gastric<br>Bypass: 197 (100) | 3.3 ± 2.6 | Patients with<br>BMI >35 kg/m<sup>2</sup> | (24) |
|  |  |  | 163<br>control |  | 3.3 ± 2.6 |  |  |
| Nguyen *et al.*, 2020 | Retrospective<br>Cohort | USA | 50,296<br>surgery | N/A | N/A | Adult patients with<br>class II (BMI 35.0<br>to 39.9 kg/m<sup>2</sup>) or<br>class III obesity<br>(BMI > 40 kg/m<sup>2</sup>) | (25) |
|  |  |  | 1,650,647<br>control | - | N/A |  |  |
| Yuan *et al.*,<br>2021 | Retrospective<br>Cohort | Minnesota,<br>USA | 308<br>surgery | Roux-en-Y Gastric<br>Bypass: 308 (100) | 4.6 (2.7–7.1) | Patients with<br>class II-III obesity<br>(BMI > 35 kg/m<sup>2</sup>) | (26) |
|  |  |  | 701<br>control | - | 4.6 (2.7–7.1) |  |  |

**Table 2.**
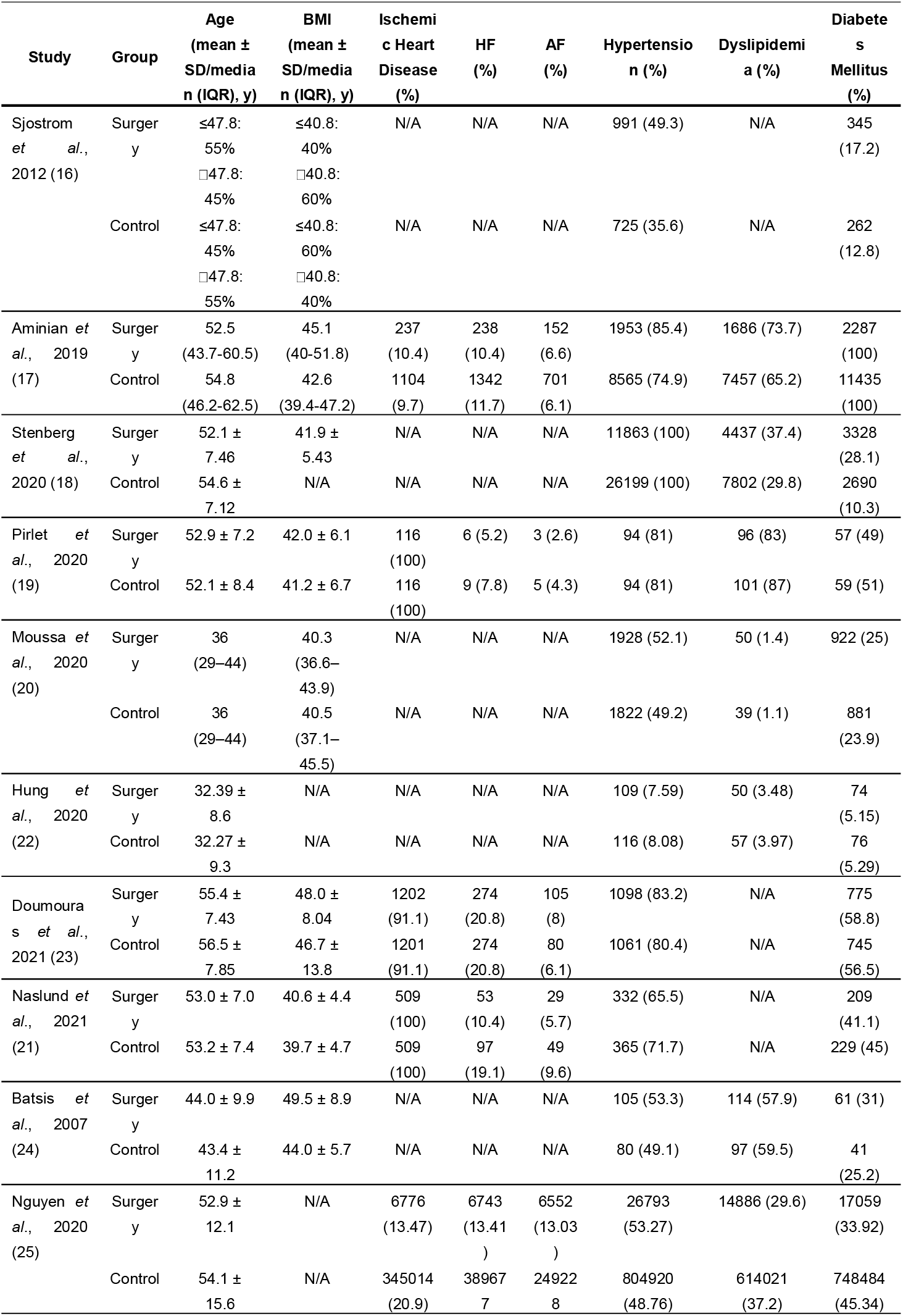
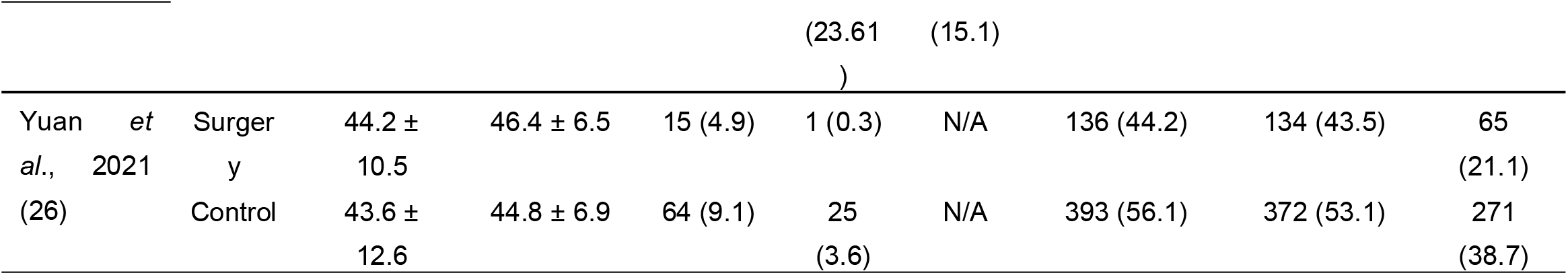
Characteristics of study population.

### 3.2 Detailed Descriptions of the Included Studies

The study by Sjostrom *et al*. (16) was conducted on 2,010 subjects underwent bariatric surgery and 2,037 subjects receiving conservative managements. The selected patients were within the range of 37 to 60 years old, had a BMI of at least 34 kg/m^2^ for men and 38 kg/m^2^ for women. Patients with earlier surgical operation for peptic ulcer, earlier bariatric surgery, history of gastric ulcer within the last 6 months, malignancy, MI within the last 6 months, bulimic eating pattern, drug or alcohol abuse, and psychiatric problems contraindicating surgery were excluded from the study. The median follow-up of the study was 14.7 years. In the study, 199 CV events (both fatal and non-fatal) in the bariatric surgery group and 234 events in the control group were reported (unadjusted HR = 0.83; 95% CI 0.69-1.00, *p*=0.05). Analyses of incidence of fatal and non-fatal MI and stroke were also lower in the surgery group (fatal MI [HR = 0.52; 95% CI 0.31-0.89; *p*=0.02] and total MI [HR = 0.71; 95% CI 0.54-0.94; *p*=0.02]; fatal stroke [HR = 0.34; 95% CI 0.12-1.00; *p*=0.05] and total stroke [HR = 0.66; 95% CI 0.49-0.90; *p*=0.008]). Of note, the baseline plasma insulin was higher in the bariatric surgery group, suggesting an improvement of glucose metabolism.

In the study conducted by Aminian *et al*. (17), a total of 2,287 and 11,435 subjects were included in the surgery group and in the control group, respectively. The study had a median follow-up of 3.9 years. The patients’ selection was based on inclusion criteria of age between 18 to 80 years, BMI of 30 kg/m^2^ or greater, and a diagnosis of diabetes (either glycated hemoglobin [HbA1c] level of ≥6.5% or the consumption of diabetes medications). Patients with history of solid organ transplant, severe HF (ejection fraction <20%), malignancy or peptic ulcer were excluded. Primary endpoints of the study (i.e., the composite of first occurrence of all-cause mortality, coronary artery events, cerebrovascular events, HF, nephropathy and AF) were observed in 385 patients within the bariatric surgery group and 3,243 patients in the control group with a cumulative incidence of primary endpoint at 8-year follow-up of 30.8% in the surgical group and 47.7% in the control group (adjusted HR = 0.61; 95% CI 0.55-0.69; *p*<0.001). Further analysis also showed that subjects in the surgery group had lower all-cause mortality (HR = 0.59; 95% CI 0.48-0.72, *p*<0.001), HF (HR = 0.38; 95% CI 0.30-0.49; *p*<0.001), coronary artery disease (HR = 0.69; 95% CI 0.54-0.87; *p*=0.002), cerebrovascular disease (HR = 0.67; 95% CI 0.48-0.94; *p*=0.02), nephropathy (HR = 0.40; 95% CI 0.31-0.52; *p*<0.001) and AF (HR = 0.78; 95% CI 0.62-0.97; *p*=0.03) than the control group. The bariatric surgery group was also associated with better weight and HbA1c reductions.

Stenberg *et al*. (18) conducted a matched cohort study involving 11,863 subjects in the surgery group and 26,199 subjects in the control group. The study was performed on subjects with age >18 years old, obesity and a history of hypertension. The incidences of MACE (the first occurrence of acute coronary syndrome, cerebrovascular event, fatal CV event or unattended sudden cardiac death) were reported in 379 subjects in the surgery group and 1,125 subjects in the control group (unadjusted HR = 0.73; 95% CI 0.65–0.82; *p*<0.001). Individual analysis of MACE showed that the surgery group had a significantly reduced risk of acute coronary syndrome (adjusted HR = 0.53; 95% CI 0.42–0.67; *p*<0.001) with no significance in cerebrovascular events (adjusted HR = 0.81; 95% CI 0.66–1.01; *p*=0.063). Improvements of hypertension, diabetes and dyslipidemia were also documented in this study.

The study conducted by Pirlet *et al*. (19) included 116 subjects in both surgery and control groups. The study was conducted on subjects who were obese and had stable coronary artery disease. The study showed that in the surgery group, the incidence of all-cause mortality and CV events were significantly lower than in the control group (HR = 0.55; 95% CI 0.30–0.98; *p*=0.044 and HR = 0.64; 95% CI 0.41– 0.99; *p*=0.046, respectively). The bariatric surgery group also had a better weight reduction than control (−28.1±20.5 vs. -3.7±14.4; *p*<0.00001). Next, the study by Moussa *et al*. (20) opted 7,402 subjects, divided equally between surgery and control groups. The study included subjects with BMI >35kg/m^2^ without any history of previous MACE. In the study, the bariatric surgery group had significantly lower fatal and non-fatal cardiac events (HR = 0.41; 95% CI 0.274–0.615; *p*<0.001). Moreover, bariatric surgery was also correlated with a better weight reduction, a lower incidence of MI (HR = 0.412; 95% CI 0.280–0.606; *p*<0.001) and a higher diabetes resolution (HR = 3.97; 95% CI 3.20–4.93; *p*< 0.001). Naslund *et al*. (21) also conducted a study on obese patients with a history of previous MI. In this study of 566 patients, the bariatric surgery group had lower MACE (HR = 0.44; 95% CI 0.32– 0.61), MI (HR 0.24; 95% CI 0.14–0.41) and new onset HF during follow up.

Next, Hung *et al*. (22) conducted a study on 2,872 subjects (1,436 individuals in each group) within 18–55 years old who had attempted conservative methods, had a BMI >35 kg/m^2^ with comorbidities or >40 kg/m^2^ and had no psychiatric disorders (e.g., major depression, anxiety or bulimia nervosa). The primary endpoint was hospitalization due to MI, intracranial hemorrhage, ischemic stroke or transient ischemic attack. As the result, the bariatric surgery group had significantly lower total CV events (HR = 0.168; 95% CI 0.085–0.328; *p*< 0.001), risk of MI (HR = 0.186; 95% CI 0.054–0.643; *p*=0.008) and cerebrovascular events (HR = 0.162; 95% CI 0.073–0.360; *p*<0.001).

In the study by Doumouras *et al*. (23), a total of 2,638 subjects were included, with 1,319 individuals in each group. The study was conducted on subjects with obesity (BMI >35 kg/m^2^) and a history of any CVD (e.g., ischemic heart disease or HF) with exclusions of age ≥70 years, malignancy, active substance use, pregnancy, previous solid organ (lung, liver or heart) transplant and severe liver disease with ascites. After a median follow-up of 4.9 years, the surgery group was shown to have lower MACE occurrence (HR = 0.58; 95% CI 0.48–0.71; *p*<0.001) and incidence of MI (HR = 0.63; 95% CI 0.42–0.96; *p*=0.03) than the control group. Next, Batsis *et al*. (24) conducted a study on 197 obese subjects (BMI >35 kg/m^2^) with a history of Roux-en-Y gastric bypass and 163 subjects without surgery, and reported 15 cases of MACE, 6 cases in surgery group and 9 cases in control group, although the statistical significance was not reached.

Nguyen *et al*. (25) employed data of 1,700,943 subjects from the 2012-2016 United States National Inpatient Sample (NIS) of the Healthcare Cost and Utilization Project (HCUP). A total of 1,650,647 subjects were included in the control group and 50,296 subjects in the surgery group. They showed that the surgery group had lower MACE (6.71% vs. 13.86%; *p*<0.001), MI (1.31% vs. 2.82%; *p*<0.001), ischemic stroke (0.33% vs. 0.44%; *p*<0.001) and HF (0.84% vs. 1.78%; *p*<0.001).

Finally, the study by Yuan *et al*. (26) included 308 subjects underwent Roux-en-Y gastric bypass and 701 subjects in the control group. The study was conducted on obese patients with BMI >35kg/m^2^ and reported that Roux-en-Y gastric bypass surgery yielded lower MACE (adjusted HR = 0.62; 95% CI 0.44–0.88; *p*=0.008) and mortality (adjusted HR = 0.51; 95% CI, 0.26–0.96; *p*=0.04) than control. Moreover, metabolic profiles (i.e., hypertension, diabetes and dyslipidemia) of the subjects were also improved by bariatric surgery.

### 3.3. The Incidence of MACE

Out of 11 studies included in the review, one study by Yuan *et al*. (26) was excluded from the meta-analysis because the original data of the incidence of MACE was not reported. From remaining 10 studies, the incidence of MACE was pooled in a meta-analysis, and a total of 4,720 cases of MACE were found in bariatric surgery group and 234,199 cases in no-surgery group. The heterogeneity test revealed that the studies were heterogenous (*I*^*2*^= 93%), therefore a random-effect model was used in the meta-analysis. As the result, the meta-analysis showed that there was a significant reduction of MACE in bariatric surgery group compared to no-surgery group (OR = 0.49; 95% CI 0.40-0.60; *p*<0.00001; *I*^*2*^= 93%). The meta-analysis was presented as a forest plot and displayed in **Figure 4**.

**Figure 4.**
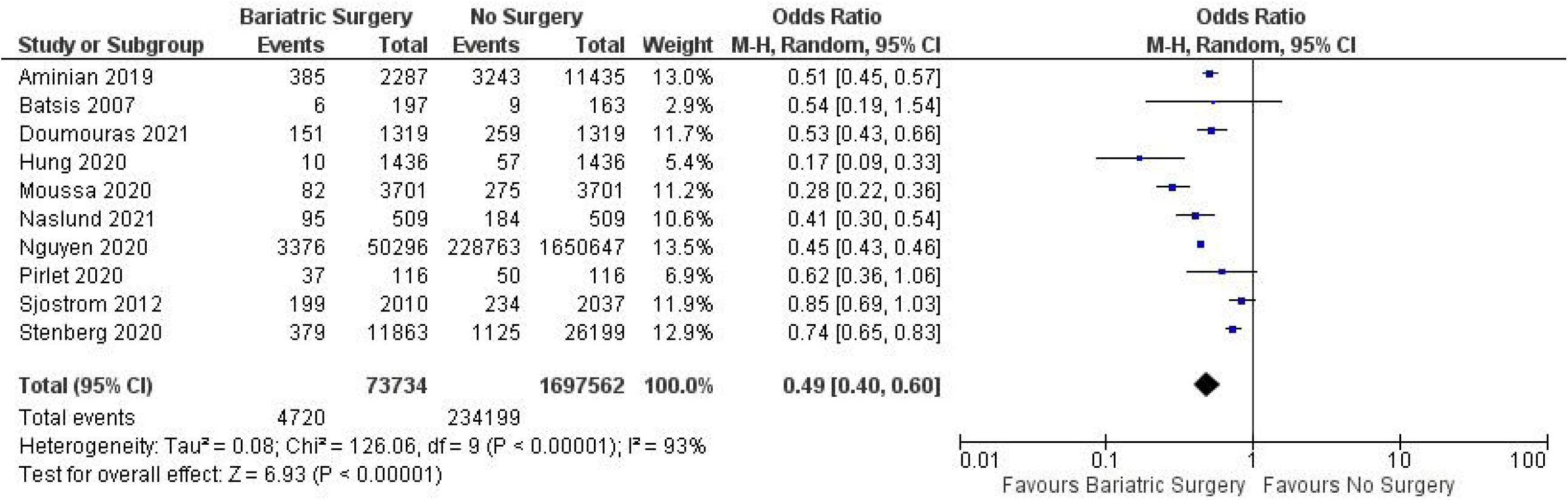
Forest Plot of MACE incidence comparing bariatric surgery with no surgery.

### 3.4. Publication Bias

The publication bias was assessed visually using the Begg’s funnel plot. As depicted in **Figure 5**, there was no asymmetry in the funnel plot, indicating the absence of apparent publication bias in the study.

**Figure 5.**
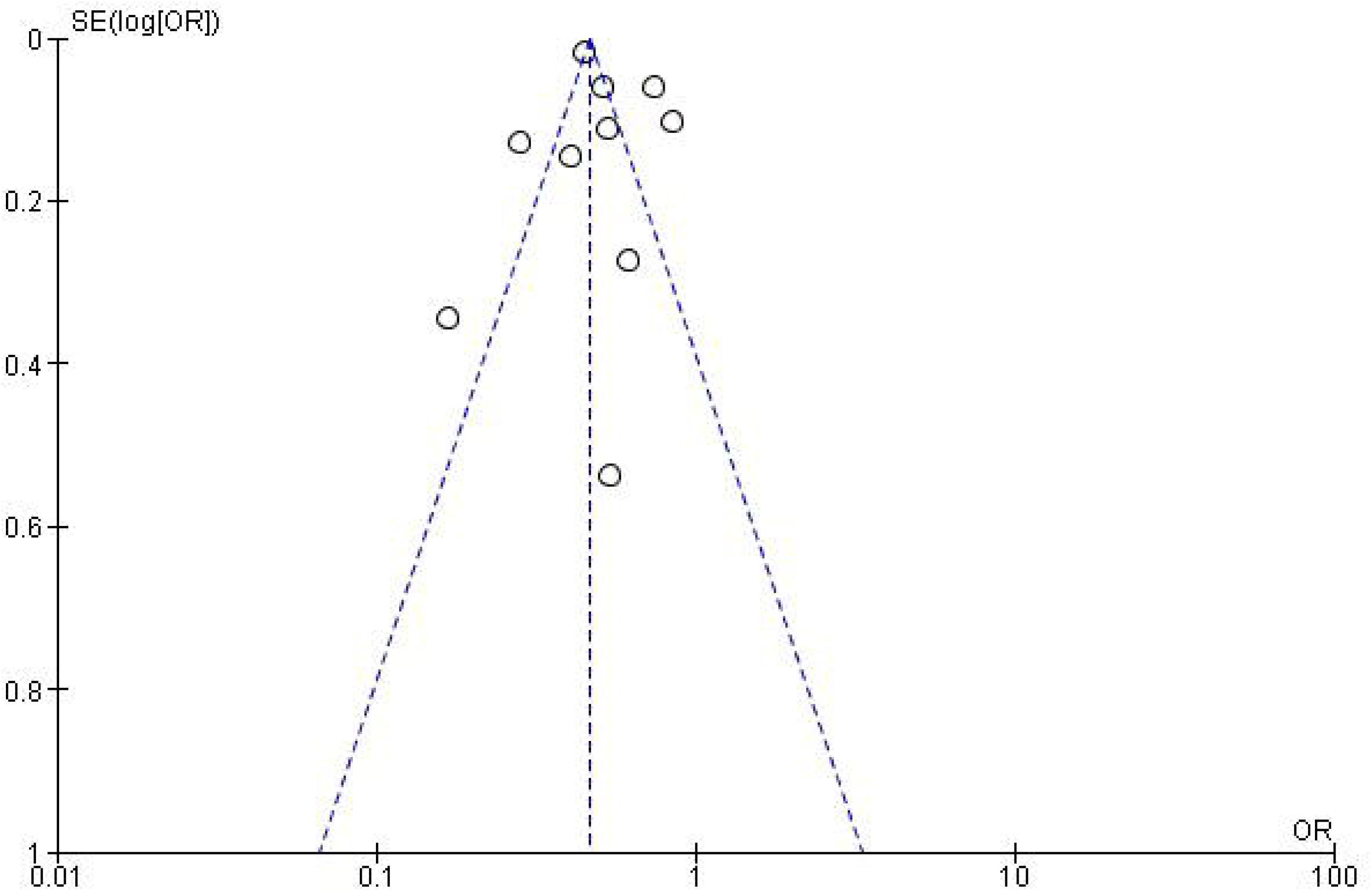
Begg’s funnel plot for publication bias assessment.

## 4. Discussion

Obesity is a notable risk factor for CV and metabolic diseases, and it has been shown to increase the risk for coronary artery disease, HF, cardiac arrhythmias and diabetes mellitus (3, 27). Despite the established association between obesity and high CV risk, in recent years, several studies reported the presence of “obesity paradox” (10-12), a phenomenon in which obesity lowers the MACE and shows a better prognosis as compared to underweighted or normal-weighted patients. At present, this notion is still unclear and debatable (28), and the (patho)physiology behind “obesity paradox” has not been fully elucidated. This paradox leads to a question whether a significant weight reduction in obese population is advantageous to lower the incidence of MACE. The objective of our study was to evaluate the efficacy of bariatric surgery on the reduction of MACE in obese patients with CVDs and our meta-analysis showed that obese individuals who underwent bariatric surgery had less occurrence of MACE as compared to the obese subjects with no surgery (OR = 0.49; 95% CI 0.40-0.60). Interestingly, this finding was consistently observed across studies, strongly indicating that bariatric surgery, and presumably a significant weight reduction in general, improved the overall CV outcomes in such specific population.

Mechanistically, there are several mechanisms through which excess adiposity could alter the body homeostasis (e.g., cellular metabolism and CV physiology). Excess adiposity induces the expression of protein tyrosine phosphatases (PTP), such as PTP-1B and leukocyte antigen-related phosphatase (LAR), which were shown to dephosphorylate the insulin receptor and insulin receptor substrate-1 (IRS-1) *in vitro*, resulted in the derangement of insulin sensitivity and energy homeostasis (29). Adipose tissue is also an endogenous source of several proinflammatory cytokines, such as tumor necrosis factor (TNF)-α, interleukin (IL)-1, IL-6, plasminogen activator inhibitor-1 (PAI-1), C-reactive protein (CRP) and monocyte chemoattractant protein-1 (MCP-1). More recently, the possible involvement of NOD-, LRR- and pyrin domain-containing protein-3 (NLRP3) inflammasome in obesity has also been articulated (30). Additionally, excess adiposity induces a high leptin level and consequently activates nicotinamide adenine dinucleotide phosphate (NADPH) oxidases (NOX) and induces oxidative stress (31). Such proinflammatory nature of adipose tissue leads to higher risks of inflammation, thrombosis and insulin resistance (29). Excess adiposity may also activate RAAS, promoting salt and water retention, and vasoconstriction. These mechanisms together with the obesity-induced autonomic nervous system remodeling could facilitate hypertension, cardiac arrhythmias (e.g., AF) and structural remodeling of the heart (i.e., HF). Additionally, excess adiposity could stimulate myocardial fat deposition (3). Subsequently, obesity-induced insulin resistance may lead to type 2 diabetes mellitus, another notable risk factor for CVDs through hyperglycemia, atherosclerotic plaque formation and diabetes-induced vasculopathy (32).

Weight reduction, either through surgical or non-surgical approach, has been shown to have positive effects on CV physiology. A study by Haase and colleagues (33) reported that a 13% reduction of body weight in obese patients significantly reduced CVD risk, including diabetes, hypertension and dyslipidemia. Moreover, studies also showed that obese patients who lost their weight had lower systolic blood pressure, HbA1c, low-density lipoprotein (LDL), triglycerides and CRP, and a higher high-density lipoprotein (HDL) (33, 34). Non-surgical approaches, such as lifestyle/diet modifications and pharmacological intervention, are the primary managements of obesity. However, studies reported that such non-surgical approaches had limited effectivity. For example, lifestyle modifications only yielded around 10% of weight loss in one year. Additionally, only 5.3% of the subjects could maintain the attained weight loss within 8 years of observation, and the remainders regained their weight (6, 35). Therefore, surgical approach (i.e., bariatric surgery) is highly considered in particular situation in which the non-surgical approaches fail to reach the weight loss target.

Bariatric or metabolic surgery encompasses any means of surgical approaches to induce weight loss. It is indicated in patients with BMI >40 kg/m^2^ or BMI >35 kg/m^2^ with comorbidities (e.g., CVDs and diabetes mellitus) (36). There are several common procedures in bariatric surgery, such as restrictive (e.g., vertical banded gastroplasty, gastric banding and sleeve gastrectomy) and malabsorptive procedures (e.g., jejunoileal bypass, duodenal switch and Roux-en-Y gastric bypass). Several procedures (e.g., jejunoileal bypass, vertical banded gastroplasty and gastric banding) are less frequent due to undesirable adverse effects, high rates of complications or reoperation, and low efficacy for the long term (36, 37). Bariatric surgery has been reported to improve CV outcomes via the improvements of CV risk factors (e.g., hypertension, diabetes and dyslipidemia) and cardiac function. Several studies have reported the benefits of bariatric surgery in glucose and fat metabolisms. For example, bariatric surgery improved diabetes mellitus through the improvement of insulin sensitivity. Bariatric surgery (i.e., Roux-en-Y gastric bypass) increased the release of postprandial glucagon-like peptide-1 (GLP-1), thus increasing insulin secretion. Additionally, the sudden negative-calorie balance post-surgery induced a normalization of blood glucose within days after surgery (37). Another study also demonstrated that bariatric surgery increased HDL cholesterol and lowered both LDL cholesterol and triglycerides (38), although the exact mechanism remains unknown. Several improvements of cardiac function were also observed in obese patients after bariatric surgery, including the reduction of left ventricular mass and the improvement of left ventricular ejection fraction (39). On a whole, these effects result in the betterment of overall metabolic and CV functions, and subsequently reducing the occurrence of MACE, as observed in our meta-analysis. Nonetheless, there are several complications of bariatric surgery, depending on the type of procedures. In general, risks of gastrointestinal obstructions (e.g., stenosis of anastomosis, intussusception and internal hernia) cannot be omitted. Specifically, in gastric banding, there is a risk for gastric necrosis. Additionally, dumping syndrome (i.e., combinations of sweating, dizziness, palpitations, abdominal pain, nausea, vomiting and/or diarrhea due to rapid gastric emptying) could also occur following bariatric surgery (40).

At last, there are several limitations of this systematic review/meta-analysis. First, our study did not differentiate outcomes based on different bariatric procedures because of the insufficiency of available data. Second, the unavailability of RCT also limits the interpretability of our finding. In the future, RCTs would be required to support/confirm our finding. Additionally, further analysis based on individual type of bariatric procedures and a more detailed analysis on individual MACE components are warranted.

## 5. Conclusions

Our systematic review/meta-analysis highlighted a significantly lower MACE in obese patients with CVDs underwent bariatric surgery as compared to patients with no surgery. Such MACE-lowering effect could be due to the reduction of CV risk/burden, through the normalization of glucose and fat metabolisms, the improvement of cardiac function and the improvement of overall CV outcomes.

## Data Availability

The data that support the findings of this study are available upon reasonable request.

## Acknowledgement

None

## Author Contributions

Conceptualization, methodology, formal analysis, investigation, resources, data curation, writing—original draft preparation, writing—review and editing, visualization, A.S. and H.S. All authors have read and agreed to the published version of the manuscript.

## Funding

None

## Notes

**Disclosure statement:** the authors have no conflict of interest to declare.

### Competing Interest Statement

The authors have declared no competing interest.

### Author Declarations

This is a meta-analysis. Ethical approval is not required.

